# Diet and physical activity during the COVID-19 lockdown period (March-May 2020): results from the French NutriNet-Santé cohort study

**DOI:** 10.1101/2020.06.04.20121855

**Authors:** Mélanie Deschasaux-Tanguy, Nathalie Druesne-Pecollo, Younes Esseddik, Fabien Szabo de Edelenyi, Benjamin Allès, Valentina A. Andreeva, Julia Baudry, Hélène Charreire, Valérie Deschamps, Manon Egnell, Leopold K. Fezeu, Pilar Galan, Chantal Julia, Emmanuelle Kesse-Guyot, Paule Latino-Martel, Jean-Michel Oppert, Sandrine Péneau, Charlotte Verdot, Serge Hercberg, Mathilde Touvier

**Affiliations:** Sorbonne Paris Nord University, Inserm, Inrae, Cnam, Nutritional Epidemiology Research Team (EREN), Epidemiology and Statistics Research Center – University of Paris (CRESS), Bobigny, France; Paris-Est University, Lab’Urba, UPEC, Créteil, France; Nutritional Surveillance and Epidemiology Team (ESEN), French Public Health Agency, Sorbonne Paris Nord University, Epidemiology and Statistics Research Center – University of Paris (CRESS), Bobigny, France.; Department of Public Health, Avicenne Hospital, Bobigny, France; Department of Nutrition, Institute of Cardiometabolism and Nutrition, Sorbonne University, Pitié-Salpêtrière Hospital, Paris, France. France

## Abstract

**Background:** Since December 2019, the coronavirus disease (COVID-19) has massively spread, with overwhelming of health care systems and numerous deaths worldwide. To remedy this, several countries, including France, have taken strict lockdown measures, requiring the closure of all but essential places. This unprecedented disruption of daily routines has a strong potential for disrupting nutritional behaviours. Nutrition being one of the main modifiable risk factors for chronic disease risk, this may have further consequences for public health. Our objective was therefore to describe nutritional behaviours during the lockdown period and to put them in light of individual characteristics.

**Methods:** 37,252 French adults from the web-based NutriNet-Santé cohort filled lockdown-specific questionnaires in April-May 2020 (nutritional behaviours, body weight, physical activity, 24h-dietary records). Nutritional behaviours were compared before and during lockdown using Student paired t-tests and associated to individual characteristics using multivariable logistic or linear regression models. Clusters of nutritional behaviours were derived from multiple correspondence analysis and ascending hierarchical classification.

**Results:** During the lockdown, trends for unfavourable nutritional behaviours were observed: weight gain (for 35%; +1.8kg on average), decreased physical activity (53%), increased sedentary time (63%), increased snacking, decreased consumption of fresh food products (especially fruit and fish), increased consumption of sweets, biscuits and cakes. Yet, opposite trends were also observed: weight loss (for 23%, −2kg on average), increased home-made cooking (40%), increased physical activity (19%). These behavioural trends related to sociodemographic and economic position, professional situation during the lockdown (teleworking or not), initial weight status, having children at home, anxiety and depressive symptoms, as well as diet quality before the lockdown. Modifications of nutritional practices mainly related to routine change, food supply, emotional reasons but also to voluntary changes to adjust to the current situation.

**Conclusion:** These results suggest that the lockdown led, in a substantial part of the population, to unhealthy nutritional behaviours that, if maintained in the long term, may increase the nutrition-related burden of disease and also impact immunity. Yet, the lockdown situation also created an opportunity for some people to improve their nutritional behaviours, with high stakes to understand the leverages to put these on a long-term footing.

## INTRODUCTION

The world is currently facing a major pandemic of the coronavirus disease (COVID-19) caused by the severe acute respiratory syndrome coronavirus 2 (SARS CoV-2). First reported in Wuhan, China, in December 2019, the outbreak of COVID-19 has since massively spread all over the globe, counting as of June 3, 2020, more than 6,348,900 confirmed cases and 380,800 deaths [1,2]. Considering the numerous unknowns surrounding the SARS CoV-2, the absence of treatment, and in order to stop a fast-growing transmission of the disease and the subsequent overwhelming of hospitals and health care systems, several countries have opted for strict lockdown measures.

In France, with over 151,600 confirmed cases of COVID-19 and 29,000 deaths reported as of June 3, 2020, such lockdown measures were declared by the government on March 17^th^, requiring the closure of all but essential public places, businesses and services, including schools and universities, workplaces, non-food shops, open and green spaces, recreational spaces (including sports clubs) and local (outdoor) food markets (when physical distancing could not be applied). The population was required to stay home and only go outside in close proximity for essential needs (including grocery shopping, medical care, legal obligations or limited recreational physical activity within a 1km radius from home), under police control. Only workers from essential sectors (e.g., healthcare, food and drug manufactures and supplies, garbage collection, city cleaning, vehicle and technology maintenance) were allowed to keep their usual activity, with protective equipment and physical distancing guidelines. As a result, a vast majority of the population either were asked to telework from home or became partially unemployed, and parents had to relay school teachers at home. On May 11^th^ and then June 2^nd^ the lockdown measures were partially lifted, but the situation has not returned to “normal” yet: e.g., most employees are encouraged to keep working from home; physical distancing is required in public transports, schools, companies, shops, bars and restaurants (when open), limiting their capacity; free movement across borders is still very limited.

This unprecedented situation has left people with a sudden disruption of their daily routines, with nonetheless a variety of circumstances according to the sociodemographic and economic status and the residential area, accompanied by major uncertainties and worry/stress related to a pandemic with continuous media coverage, but also to the current (and future) professional and familial organization. The consequences of the lockdown measures for the physical and mental health of the population have therefore come into question.

In particular, the lockdown has a strong potential for disrupting nutritional behaviours, notably due to modified access to food, stopping daily travels and mobility, sedentary lifestyle, food/meals contributing to give rhythm to the day, buying food and limited physical activity being the only authorized activities outside home for most people. Nutrition (diet, physical activity, weight status) is one of the main modifiable factors for chronic disease risk, such as cardiovascular diseases, type 2 diabetes and some cancers [3,4]. Besides, mounting evidence shows the impact of nutritional factors on the functioning of the immune system through various mechanisms, including gut microbiota modulation [5,6]. Thus, nutrition may directly impact the risk of SARS CoV-2 infection and its prognosis [7–10]. It is therefore important to inform what happened during the lockdown period in terms of nutritional behaviours, and assess the related potential consequences for public health, now and in the future. Indeed, the duration of the initial lockdown period might have been sufficient for modified habits to settle in some individuals [11,12]. Moreover, the situation is not resolved yet (no vaccines or treatment to date) and new lockdown periods may occur in the future at the individual or population levels, regionally or nationally.

The NutriNet-Santé web platform offered the unique opportunity to collect a large amount of nutritional, behavioural and health data during the lockdown for > 37,000 French adults using online questionnaires and validated dietary records. The aim of the present study was to characterise nutritional behaviours in this large population of volunteers during the lockdown period and how these differed from usual nutritional behaviours, in light of individual characteristics.

## METHODS

### Study population: the NutriNet-Santé cohort

The NutriNet-Santé cohort started in France in 2009 with the objective to study the associations between nutrition and health, as well as the determinants of nutritional behaviours [13]. Recruitment of participants (adults and, more recently, teenagers aged > 15 years old) is still ongoing. The study uses a secured online platform to reach out to participants and collect data. This flexible platform allows for the rapid implementation of new research protocols as needed. The NutriNet-Santé study is conducted in accordance with the Declaration of Helsinki, and all procedures were approved by the Institutional Review Board of the French Institute for Health and Medical Research (IRB Inserm 0000388FWA00005831) and by the Commission Nationale de l’Informatique et des Libertés (CNIL 908,450 and 909,216). All participants provided informed consent with an electronic signature; this study is registered in ClinicalTrials.gov (NCT03335644).

### Data collection in the NutriNet-Santé cohort

Upon inclusion, NutriNet-Santé participants are asked to complete a set of five validated self-administered web-based questionnaires related to 1) sociodemographic and lifestyle characteristics [14], 2) health status, 3) dietary intakes [15–17], 4) physical activity (short form of the International Physical Activity Questionnaire, IPAQ [18]), and 5) anthropometrics [19,20]. These questionnaires are then repeated every 6 to 12 months during follow-up.

In particular, dietary intakes are assessed every 6 months as part of the usual cohort follow-up using three non-consecutive 24h dietary records randomly assigned over two weeks and including two weekdays and one weekend day. These web-based 24h dietary records have been validated against dietary records filled during an interview with a dietitian and against biomarkers [15–17]. Portion sizes are estimated using validated photographs, standard containers or directly in g/L. Mean daily energy, alcohol and macro- and micro-nutrient intakes are estimated using a published French food composition database comprising > 3,500 food items [21]. Amounts consumed from composite dishes are estimated using French recipes validated by food and nutrition professionals. Dietary under-reporters are detected on the basis of the method proposed by Black [22]. In turn, the IPAQ is sent each year to the participants, assessing their physical activity (vigorous intensity, moderate intensity or walking) and time spent seated on an average day (outside sleeping hours) during the past 7 days. Physical activity levels for each participant are computed as Metabolic Equivalents (MET) minutes per week.

### Data collection during lockdown

In April 2020, a set of questionnaires was sent to the NutriNet-Santé participants to collect extensive data on nutritional behaviours during the lockdown: a specific questionnaire for participants to perform a qualitative assessment of changes in their dietary habits, consumption of major food groups, snacking, food supply, physical activity and sedentary lifestyle, as well as body weight (details in Supplementary Material 1); a series of three 24h dietary records randomly assigned over two weeks during the strict lockdown period (two weekdays and one weekend day); the IPAQ (allowing the computation of MET-min/week). The 24h dietary records and the IPAQ were the same as those that are regularly sent to the participants as part of their follow-up (described above).

Body weights at the beginning and at the end of the strict lockdown period were collected from self-measures reported as integer values in questionnaires sent respectively in early April and early May 2020. In these questionnaires, participants were also asked about their weight just before the lockdown. Participants were asked if they had been able to measure their weight with a scale during this period and provide measured data. The web-based self-reported body weight has been validated in the cohort by comparison with standardized clinical measurements [19]. Body mass index (BMI) was computed as weight [kilograms] / height² [meters]. The following categories of body weight status were defined: obese (BMI ≥ 30 kg/m^2^), overweight (BMI ≥ 25 to < 30 kg/m^2^), normal-weight (BMI ≥ 18.5 to < 25 kg/m^2^), underweight (BMI < 18.5 kg/m^2^).

Finally, a questionnaire assessing participants’ exposure to SARS-CoV-2, COVID-19 infection and experience of the lockdown was simultaneously sent to participants in April 2020 as part of a national multi-cohort project (*Health, Practices, Relationships and Social inequalities in the general population during the COVID-19 crisis*, SAPRIS). This questionnaire was used to derive information regarding personal characteristics during the lockdown (professional status, presence of children or grandchildren aged < 18 years at home), including scores for depressive symptoms (Patient Health Questionnaire (PHQ)-9 scale [23]) and anxiety (General Anxiety Disorder (GAD)-7 scale [24]). Details are provided in Supplementary Material 2.

### Statistical analyses

A total of 37,252 participants living in metropolitan France filled in the specific questionnaire related to nutrition during the lockdown, which served as the basis for the main analyses. Among these, participants with available data on dietary intakes before and during lockdown from 24h dietary records, physical activity and sedentary lifestyle before and during lockdown, or weight status in May 2020 are detailed using a flowchart in Supplementary Figure 1.

Indicators of diet quality were calculated based on the dietary intakes in food and nutrients reported by the participants in their 24h dietary records before and during the lockdown. The Alternative Healthy Eating Index (AHEI)-2010 score [25] was calculated as a measure of overall diet quality based on the dietary intakes of vegetables, fruit, whole grains, sugar-sweetened beverages, nuts and legumes, red/processed meat, long-chain (n-3) polyunsaturated fatty acids (PUFA), total PUFA, sodium and alcohol. Details of its computation are available in Supplementary Material 3. The percentage (by relative weight) of ultra-processed foods in the diet was defined using the NOVA classification, as previously described [26].

Self-reported data are summarised using numbers and percentages for categorical variables and mean and standard deviations for continuous variables. Student paired t-tests were used to compare values before and during the lockdown (dietary intakes: food groups, macro- and micronutrients, AHEI-2010 score and share of ultra-processed foods, physical activity in MET-min/week and sedentary lifestyle in hours). Variations of continuous variables during the lockdown compared to before the lockdown were computed as raw values and as percentage of variation ((value during lockdown – value before lockdown)/value before lockdown). Increased or decreased intakes in macro- and micronutrient were defined as variations of at least 10% comparing dietary records of participants during the lockdown to dietary records before the lockdown.

Comparisons across individual characteristics were performed using multivariable-adjusted logistic regression models (binary or multinomial) for categorical variables or ANCOVA models for variations of continuous variables. These models included the following characteristics (coding of the characteristics is detailed in Table 1): age, sex, current weight status (using weight reported for all in April 2020), smoking status, educational level, household monthly income, professional activity during the lockdown, marital status during the lockdown, presence of children or grandchildren aged < 18 years at home during the lockdown, regional residential area during the lockdown, urban or rural residential area during the lockdown, PHQ-9 score for depressive symptoms during the lockdown, GAD-7 score for anxiety during the lockdown, self-reported chronic disease, qualitative assessment of change in sedentary lifestyle and physical activity during the lockdown. For participants for whom data on usual dietary intakes before the lockdown was available (N = 27,658), the models additionally included the AHEI-2010 score and the proportion of ultra-processed foods in the diet.

**Table 1.** Characteristics of the study population after weighting (N = 37,252), NutriNet-Santé cohort, March-May 2020.

|  | % or Mean (SD) |
| --- | --- |
| **Sex** |  |
| Women | 52.3 |
| Men | 47.7 |
| **Age, years** |  |
| [18-25] | 4.4 |
| ]25-50] | 42.5 |
| ]50-65] | 26.3 |
| ]65-80] | 25.0 |
| > 80 | 1.8 |
| **Current weight status<sup>1</sup>** |  |
| Obesity | 11.05 |
| Overweight | 26.05 |
| Normal | 58.94 |
| Underweight | 3.96 |
| **Smoking status** |  |
| Never smoker | 45.1 |
| Former smoker | 40.6 |
| Current smoker | 14.2 |
| **Educational level** |  |
| < High-school degree | 17.16 |
| High-school degree | 15.16 |
| Undergraduate degree | 32.74 |
| Graduate degree or doctorate | 34.23 |
| Unknown | 0.72 |
| **Monthly income, € per household** |  |
| < 1430 | 8.5 |
| [1430 – 2700[ | 24.5 |
| 2700 to < 4800 | 39.1 |
| > 4800 | 14.5 |
| Unknown | 3.3 |
| Did not wish to answer | 10.1 |
| **Professional activity during the lockdown** |  |
| No professional activity prior to lockdown:<br>unemployed, retired, homemaker | 40.4 |
| Working outside home | 12.3 |
| Partially unemployed | 15.7 |
| Teleworking from home (fully) | 19.8 |
| Teleworking from home (partially) | 5.2 |
| Student | 3.2 |
| Other | 3.4 |
| **Marital status during the lockdown** |  |
| Never married | 16.3 |
| In a relationship | 16.3 |
| Married or registered partnership | 56.7 |
| Divorced or separated | 7.6 |
| Widowed | 3.1 |
| **Children or grandchildren aged under 18 at home during the lockdown? yes** | 24.6 |
| **Residential area (city size, inhabitants) during the lockdown** |  |
| City > 100,000 | 19.8 |
| City ≥ 20,000 to 100,000 | 21.6 |
| City < 20,000 | 23.1 |
| Rural area | 35.4 |
| **Regional residential area during the lockdown<sup>2</sup>** |  |
| Paris Basin | 15.8 |
| Centre-East | 12.9 |
| East | 9.2 |
| Mediterranean | 13.5 |
| North | 5.7 |
| West | 14.1 |
| Paris Region | 17.4 |
| South-West | 11.3 |
| **GAD-7 (anxiety disorders)<sup>3</sup>** | 3.2 (4.0) |
| **PHQ-9 (depressive symptoms)<sup>4</sup>** | 3.8 (4.4) |
| **Chronic disease<sup>5</sup>, yes** | 27.9 |
| **Physical activity level** |  |
| Not modified | 26.2 |
| Increased | 18.6 |
| Decreased | 52.9 |
| I don't know | 2.2 |
| **Sedentary time** |  |
| Not modified | 28.4 |
| Increased | 63.3 |
| Decreased | 4.6 |
| I don't know | 3.6 |
| **Diet quality before the lockdown (N=27,658)** |  |
| AHEI-2010 score<sup>6</sup> | 50.1 (12.4) |
| Ultra-processed foods (% food weight) | 16.7 (7.8) |

<sup>1</sup> Calculated from the current weight at the time of the questionnaire reported by the participants in April, 2020
<sup>2</sup> Regional Zones for Study and Development (ZEAT) as defined by INSEE: PARIS REGION - Ile de France; PARIS BASIN - Burgundy, Centre, Champagne-Ardenne, Lower and Upper Normandy, Picardie; NORTH - Nord Pas-de-Calais; EAST - Alsace, Franche-Comté, Lorraine; WEST- Brittany, Pays de la Loire, Poitou-Charentes; SOUTH-WEST - Aquitaine, Limousin, Midi-Pyrénées; CENTRE-EAST - Auvergne, Rhône-Alpes; MEDITERRANEAN- Languedoc-Roussillon, Provence-Alpes-Côte d'Azur, Corsica.
<sup>3</sup> The GAD-7 scale ranges from 0 to 21 points and measures an increasing severity of anxiety (minimal: 0-4, mild: 5-9, moderate: 10-14, severe: 15-21).
<sup>4</sup> The PHQ-9 scale ranges from 0 to 27 points and measures an increasing severity of depressive symptoms (minimal: 0-4, mild: 5-9, moderate: 10-14, moderately severe: 15-19, severe: 20-27)
<sup>5</sup> includes diabetes, cardiovascular diseases, hypertension, dyslipidemia, cancer, liver diseases, kidney diseases, thyroid diseases, digestive disorders, gynaecological disorders, arthritis, immunity disorders
<sup>6</sup> The AHEI-2010 score ranges from 0 to 100 points and measures an increasing diet quality

A multiple correspondence analysis (MCA) was carried out to derive profiles of nutritional behaviours during the lockdown. The following aspects of nutritional behaviours during the lockdown were included: changes in weight and qualitative assessment of changes in both sedentary time and physical activity, main reasons for modifying eating habits, self-assessed diet quality, qualitative assessment of changes in consumption of fresh fruit, vegetables, fish and red meat and of potatoes, sandwiches/pizzas/savoury pies, cheese, sweets and chocolate, biscuits and cakes, alcohol, and tea, food-storing behaviour, snacking, and stress due to lacking some foods. Two dimensions were kept based on inertia decomposition and relevance/interpretability of the obtained profiles [27] (explaining 10.6% and 6.4% of the variation respectively; coordinates of nutritional behaviours along these dimensions are shown in Supplementary Table 1). An ascending hierarchical classification (AHC) was then applied on the scores of participants along these two dimensions to define clusters of participants displaying similar nutritional behaviours during the lockdown. Characteristics of participants belonging to each cluster (as dummy variables) were obtained using multivariable logistic regression models including the same set of variables as described above (except changes in sedentary time and physical activity level during lockdown, active variables in the MCA).

To allow inferences for the general French adult population, a weighting was applied to all analyses to take into account differences in sociodemographic distribution (gender, age, area of residence, occupational category) in responders compared to the entire cohort and to the French population (SAS macro %CALMAR and the 2016 national Census INSEE data).

All tests were two-sided and P < 0.05 were considered statistically significant. Analyses were computed using SAS 9.3 (SAS Institute Inc., USA).

## RESULTS

After weighting, our study population was composed of 37,252 participants (52.3% women) with a mean age of 52.1 y (SD: 16.6). Further characteristics of the participants are shown in Table 1.

### Physical activity, sedentary lifestyle, energy intake and weight change (Figure 1)

**Figure 1.**
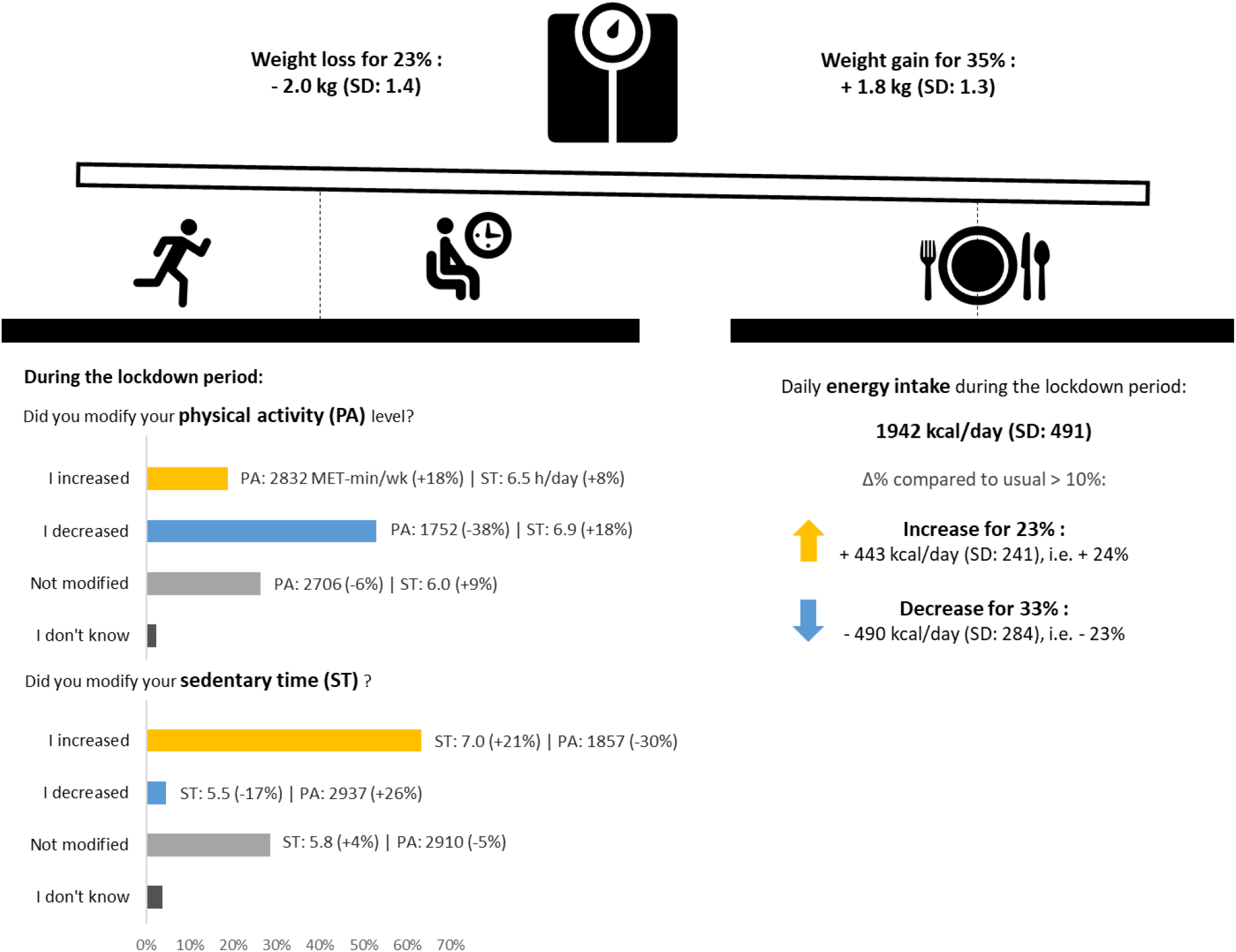
Physical activity, sedentary time, energy intake and weight variations during the lockdown. NutriNet-Santé cohort study (N = 36,917), March-May 2020. At the top: weight variations between the weight just before the lockdown and the weight in May 2020, after about 2 months of lockdown (N = 22,042); values are the mean (SD) variations in participants who gained (respectively lost) weight during the lockdown. At the bottom right, variations in daily energy intake between the usual intakes observed at the same period of the year before the lockdown and the intakes observed during the lockdown (N = 9,372); increase and decrease were defined as variations (respectively positive or negative) in energy intake of at least 10%; values are the mean (SD) variations in participants who increased (respectively decreased) their energy intake during the lockdown. At the bottom left, modifications in physical activity and sedentary time were assessed qualitatively (N = 36,917) and are shown as a bar graph; quantitative assessments of variations in physical activity levels (in MET-min/week) and sedentary time (time spent seated outside sleeping hours) between before and during the lockdown were available for subsamples (N = 29,798 for physical activity, N = 29,788 for sedentary time) and are shown next to each bar as median physical activity levels and mean sedentary time; corresponding interquartile range and standard deviation are detailed in Supplementary Table 3. Variations in quantitative values are also expressed as the % of variation compared to the value before the lockdown. Pregnant women (N = 335) were excluded from these analyses. PA, physical activity; ST, sedentary time.

A majority (52.8%) of participants reported that they decreased their level of physical activity during the lockdown (qualitative assessment). In these participants, a quantitative assessment using the IPAQ (for a subsample) highlighted median physical activity levels of 1752 MET-min/week (IQR: 742.5–3519), which is 38% less than before the lockdown (paired Student t-test: P<.0001). In contrast, a lower proportion (18.7%) reported that they increased their level of physical activity during the lockdown (median: 2832 MET-min/week, IQR: 1632–4944; +18% compared to before the lockdown; P<.0001). In addition, 63.2% of participants reported that they increased their sedentary time (qualitative assessment). In these participants, a quantitative assessment (for a subsample) highlighted an average 7.0 h/day spent seated during the lockdown (SD: 3.2), which is 21% more than before the lockdown (paired Student t-test: P<.0001). In turn, 28.5% reported no change in sedentary time (mean: 5.8 h/day spent seated, SD: 3.3; +4%; P<.0001). Participants reporting that they decreased their level of physical activity were also those reporting the longest time spent seated (mean: 6.9 h/day, SD: 3.3) and the largest increase (+18% compared to before the lockdown; P<.0001), and, similarly, participants reporting that they increased their sedentary time were also those reporting the lowest physical activity level (median: 1857 MET-min/wk, IQR: 840-3570) and the largest decrease (−30%; P<.0001).

During the lockdown, the energy intake was on average 1942 kcal/day, with 23% of participants increasing their energy intake by more than 10% (2399 kcal/day on average during lockdown, namely a 24% increase compared to before lockdown; paired Student t-test: P<.0001) and 33% of participants decreasing their energy intake by more than 10% (1664 kcal/day on average during lockdown, namely a 23% decrease; paired Student t-test: P<.0001).

A weight gain between the weight just before the lockdown and the weight in May 2020 (after about 2 months of lockdown) was observed for 35% of participants, with an average gain of 1.8 kg (SD: 1.3), and into weight loss for 23%, with an average loss of 2.0 kg (SD: 1.4). Meanwhile, the weight remained stable (no difference in reported values) for 42% of the participants. Participants who gained weight were more likely to be women, younger (aged under 25), overweight or obese (in early April), smokers, single, to have children under 18 y at home, a higher PHQ-9 score for depressive symptoms, a lower GAD-7 score for anxiety, to report having decreased their physical activity and increased their sedentary time during lockdown, and to have a usual diet before lockdown of lower nutritional quality (lower AHEI-2010 score and higher proportion of ultra-processed foods). In turn, those who lost weight were more likely to be men, aged under 50, underweight, to have an intermediate educational level (high-school or undergraduate degree), to be students, to report having increased their physical activity and decreased their sedentary time during lockdown, and to have a higher PHQ-9 score for depressive symptoms and a higher GAD-7 score for anxiety. They were also less likely to be short-time working or to work outside home during the lockdown, to have children under 18 y at home and to live in a rural area during the lockdown. Details of individual characteristics associated with weight variations are shown in Supplementary Table 2.

### Nutritional behaviours during the lockdown

#### Modifications of eating practices overall and associated reasons (Figure 2)

**Figure 2.**
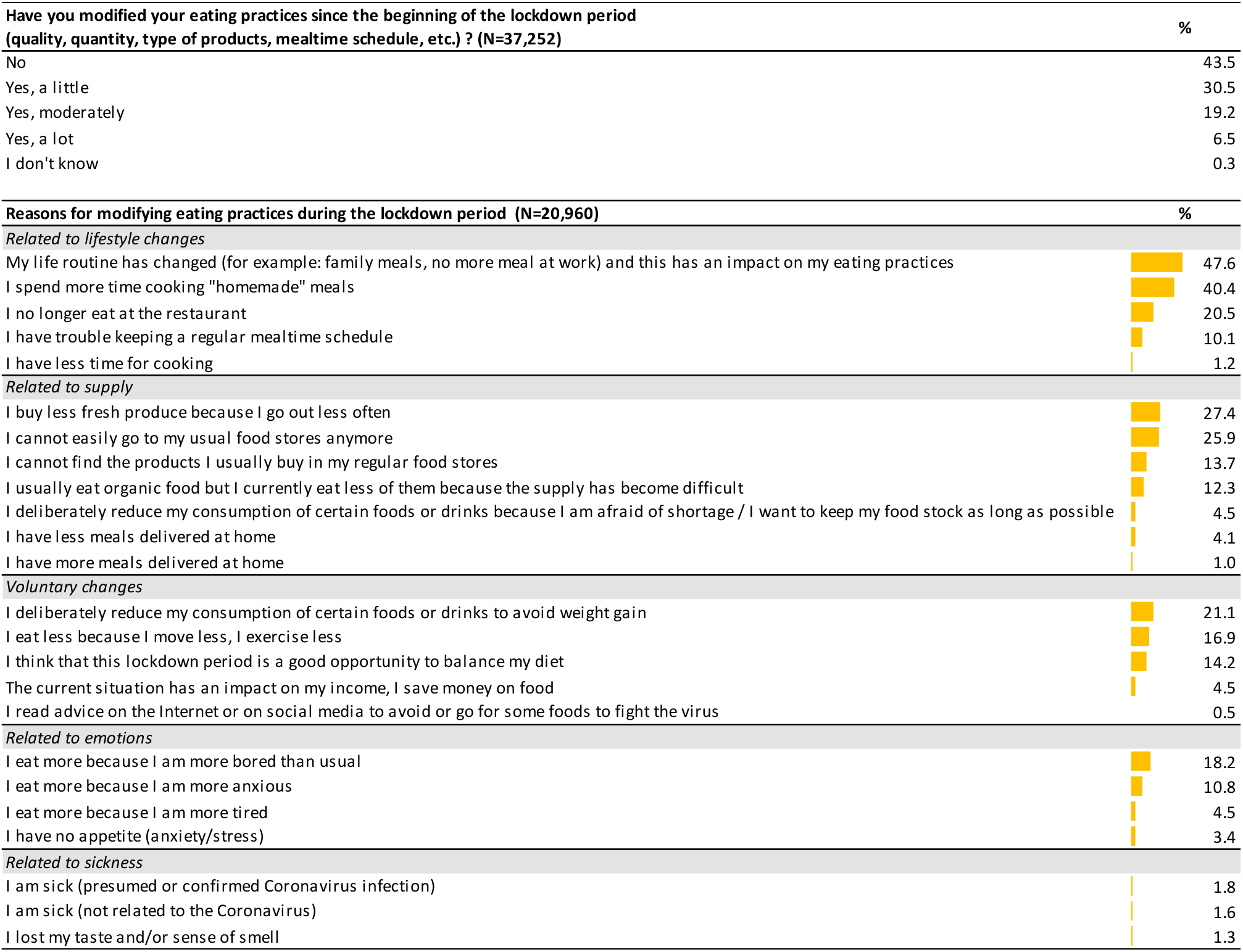
Modification of eating practices and associated reasons (several choices possible) during the lockdown, NutriNet-Santé cohort study (N = 37,252), March-May 2020. Distribution of answers to the questions “Have you modified your eating practices since the beginning of the lockdown period?” and “If yes, for what reason(s)?”. A bar graph is embedded to visualize the proportion of participants selecting each reason (multiple answers allowed), among those for answered “Yes” to the first question (N = 20,960).

Overall, 56.2% participants reported that they modified their eating practices during lockdown compared to before. The main reasons selected to explain these modifications were related to inherent lifestyle changes during the lockdown (change of routine: 47.6%, spending more time cooking “homemade” meals: 40.4%, no longer eating at the restaurant: 20.5%, trouble keeping regular mealtime schedule: 10.1%), to food supply (buying less fresh products: 27.4%, difficulty going to usual stores: 25.9%, difficulty finding usual products: 13.7%, difficulty buying organic food: 12.3%), to voluntary changes (trying to avoid weight gain: 21.1%, balancing the decrease in physical activity: 16.9%, opportunity to balance diet: 14.2%), and to emotional reasons (eating out of boredom: 18.2%, eating out of anxiety: 10.8%). Characteristics of participants associated with the main reasons for modifying eating practices are shown in Supplementary Table 4.

#### Snacking

Overall, 27.9% participants reported snacking at least once a day, every day (Figure 3). Snacking more than usual during the lockdown was reported by 21.1% of participants. Among these, 62.3% snacked at least once a day, with 18.9% at least 3 times a day. In turn, snacking less than usual was reported for 9.4% while snacking neither more nor less was reported for 69.5%. Participants who snacked more than usual were more likely to be aged under 50 y, overweight or obese, smokers, divorced or widowed, to have a higher level of education, to have children aged under 18 y at home during the lockdown, a professional activity before the lockdown (i.e. working outside home, being partially unemployed, teleworking or student during the lockdown), a higher PHQ-9 score for depressive symptoms, a lower GAD-7 score for anxiety, to report modifications (increase or decrease) in their physical activity and sedentary time and to have a usual diet before the lockdown composed of more ultra-processed foods (Supplementary Table 5).

**Figure 3.**
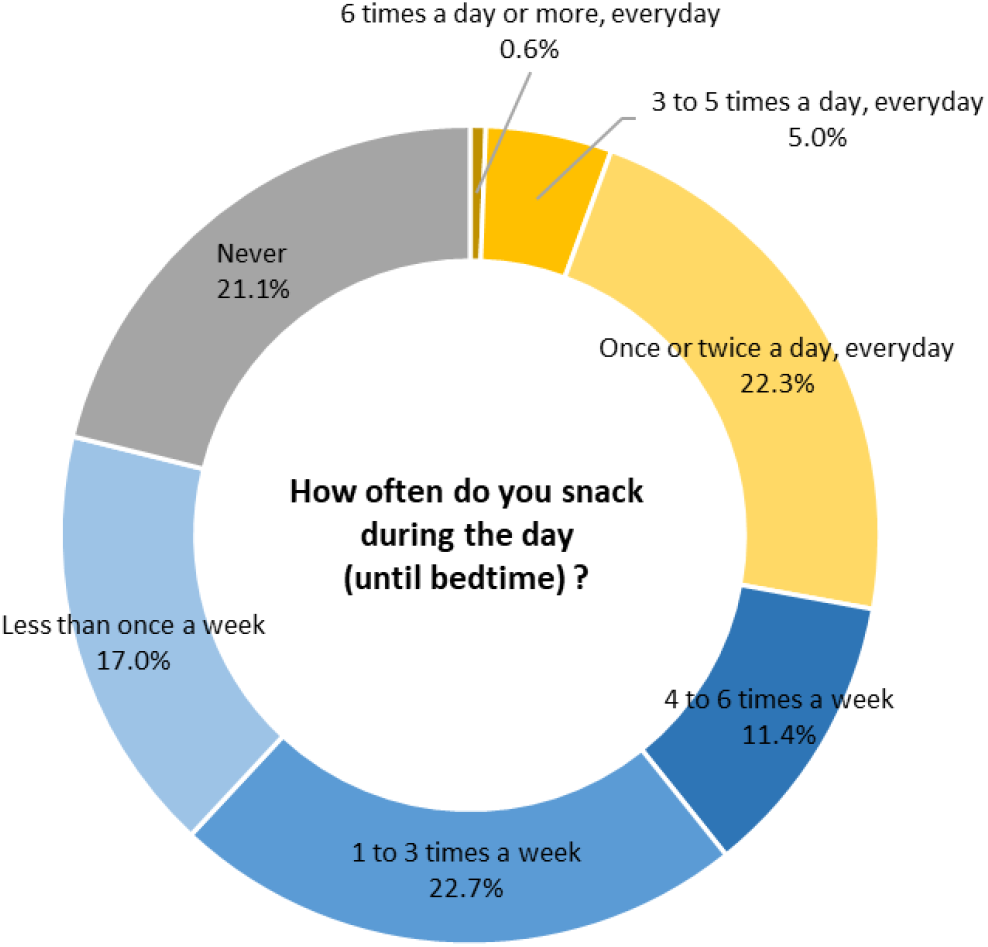
Frequency of snacking during the day during the lockdown, NutriNet-Santé cohort study (N = 37,252), March-May 2020.

#### Self-perceived diet quality during lockdown

When asked about their diet quality during lockdown compared to before, 14.1% reported that their diet was of better quality, 10.5% that their diet was of lower quality and 74.4% that their diet quality had not changed. Characteristics of participants associated with the self-perceived diet quality during lockdown are shown in Supplementary Table 5.

### Modification of food consumption during lockdown

#### Food groups

Participants were asked to qualitatively estimate how the lockdown affected their consumption of major food groups. Results in Figure 4 show an overall decrease in the consumption of fresh products: 17% of participants reported a decrease for fresh fruit and 18% for fresh vegetables, 22% for fresh red meat and 31% for fresh fish. In parallel, 14% of participants reported to have increased their consumption of frozen or canned vegetables but such proportion was much lower for frozen or canned fruit, fish or red meat. In addition, an increased consumption was reported for other products with large shelf-life, like potatoes (by 15% of the participants), legumes (15%) or nuts (12%). Other noteworthy results include an increased consumption reported for sweets and chocolate (by 22% of the participants), biscuits and cakes (20%), and cheese (18%) and a decreased consumption reported for sandwiches, pizzas or savoury pies (17%). Looking at drinks, an increased consumption of alcoholic drinks was reported by 15% of the participants (and a decrease by 12%), as well as an increase for tea (20%) and tap water (13%). These qualitative assessments were confirmed using quantitative data comparing dietary records of participants during the lockdown to dietary records before the lockdown (see Supplementary Table 6 for details).

**Figure 4.**
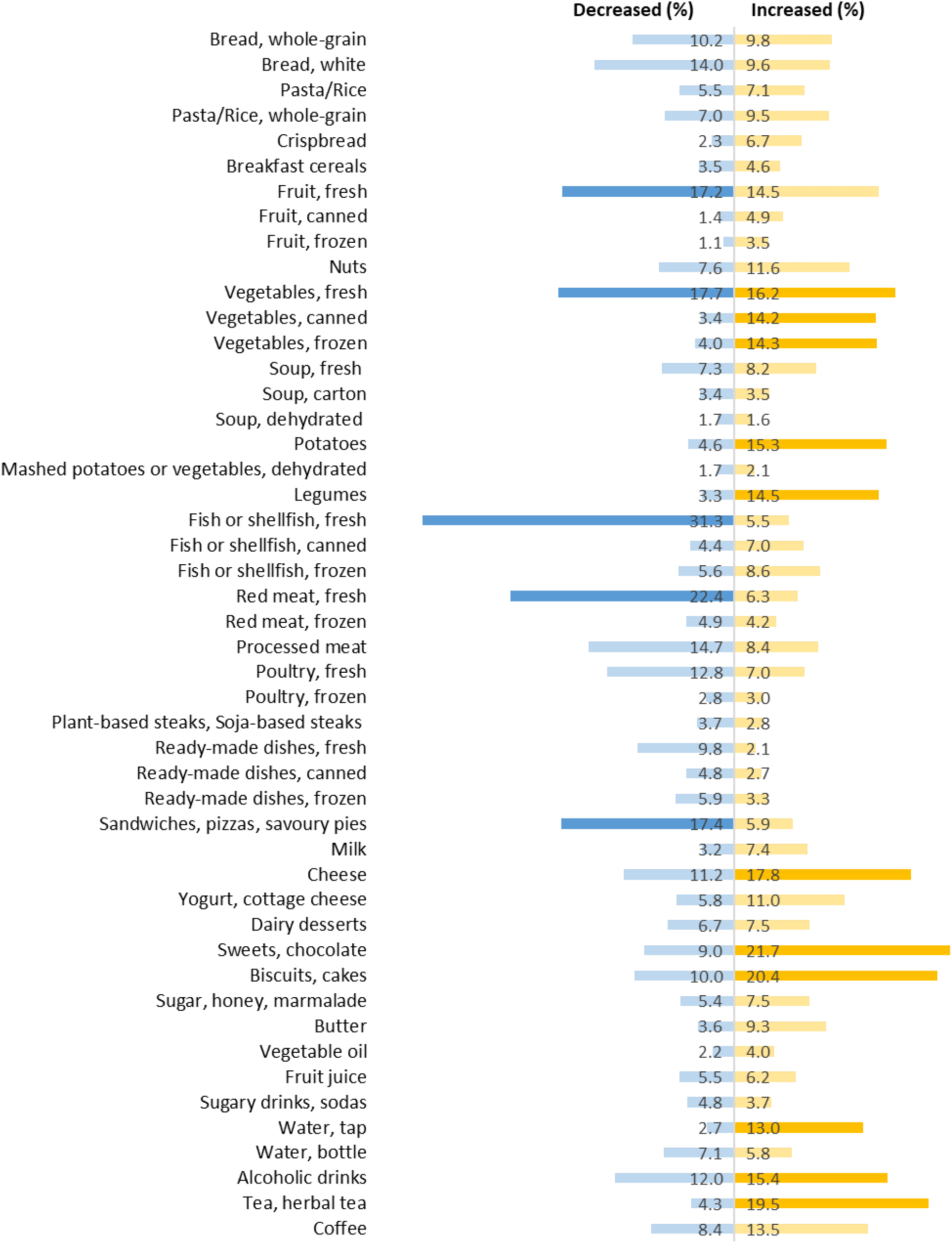
Modification of consumption of major food groups during lockdown, NutriNet-Santé cohort study (N = 37,252), March-May 2020. Bars indicate the % of participants who declared having increased or decreased the consumption of the food group of interest during lockdown (corresponding number on the respective bars); brighter colours represent % above 15% and/or a difference of % between those who increased and those who decreased of more than 10%

#### Changes in macro- and micronutrient intakes

Consistently, more than half of the participants decreased their intakes for macro- or micronutrients typically related to the intake of fish: n-3 polyunsaturated fatty acids (EPA: for 59% participants, DPA: for 61%, DHA: for 58%), iodine (for 52%), but also more generally of animal products: heme iron (for 57%), vitamin D (for 52%) and vitamin B12 (for 54%). In turn, alcohol intakes increased for 25% participants (+8g/day) and decreased for 53% (−7g/day). More details are available in Supplementary Table 7.

#### Overall diet quality (Supplementary Table 6)

As regards to the quality of diet on the whole, two opposite dietary trends were observed: 37% of the participants decreased their AHEI-2010 score by at least 10% compared to before lockdown (−13 points on average), while an increase was observed for 27% (+12 points); symmetric changes were observed for the proportion of ultra-processed foods in the diet with an increase for 42% (+6% of ultra-processed foods) and a decrease for 42% (−6%).

### Food supply

#### Stress of lacking some foods and food storing behaviour during lockdown

Overall, 27.1% of participants reported that they were somehow stressed from the idea of lacking some food during the lockdown. Only 3.3% of participants reported that they stored more food than usual at home because they were afraid of food shortages, while 45% stored more food due to a reduced frequency of grocery shopping. Characteristics of participants associated with these behaviours are shown in Supplementary Table 5.

#### Sources of food supply (Figure 5)

During the lockdown, individuals used on average 3.6 (SD: 1.7) different sources of food supply, which is 1.1 less than usual (paired Student t-test: P<.0001). The top 3 sources of supply during the lockdown were the supermarket (66%), the bakery (60.3%) and the local grocery store (41.3%), the latter owing its third place from reduced visits (compared to usual) to local (outdoor) markets (which showed the sharpest reduction, especially in cities > 20,000 inhabitants), hypermarkets, and local shops such as the butcher’s, fishmonger’s or greengrocer’s. Contrary to the reduction observed for all sources of supply, slight increases were observed for internet, phone or mail-order purchases and for purchase of food baskets from farmers or AMAP (associations supporting small farming), and only a small decrease was observed for organic food stores. Overall, 6.3% participants declared that they were not living in their usual home during the lockdown, which may have impacted the sources of supply.

**Figure 5.**
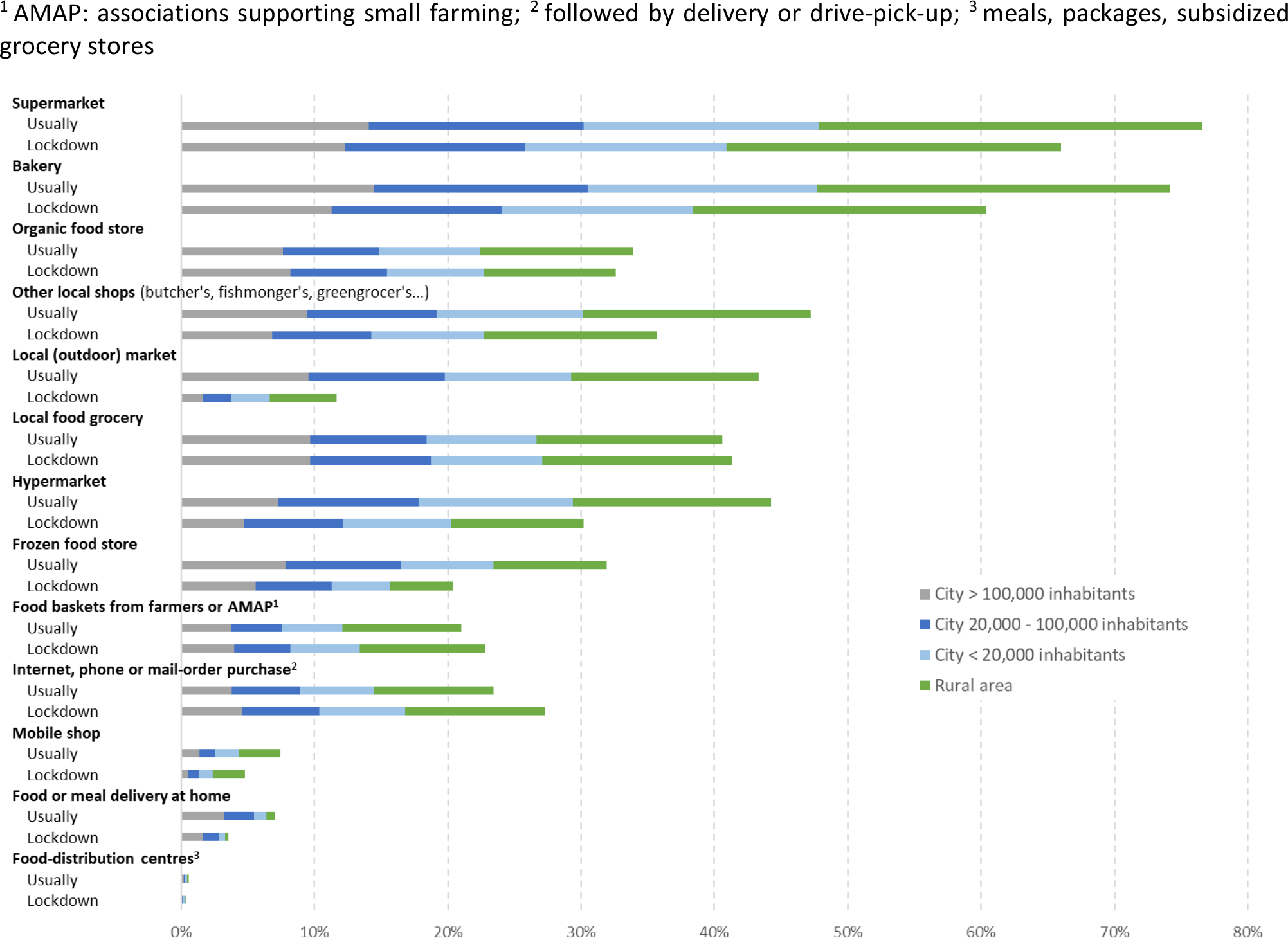
Sources of food supply usually and during lockdown, according to the urban level of the residential area during the lockdown. NutriNet-Santé cohort study (N = 37,252), March-May 2020.

### Profiles of nutritional behaviours during lockdown

Two main synthetic factors of nutritional behaviours during the lockdown were identified using MCA, and lead to identify three clusters of participants in the AHC. Cluster 1 (42.9% of participants) corresponded to participants for whom nutritional behaviours were not much impacted by the lockdown period (e.g., stable weight, physical activity, food consumption; Supplementary Table 8). This cluster was composed of participants more likely to be older, men, normal-weight, current smokers, with a lower level of education, in a relationship (married or not), living in cities with < 100,000 inhabitants or rural areas, with lower scores on the GAD-7 scale for anxiety and the PHQ-9 scale for depressive symptoms, and with a higher usual diet quality before the lockdown period (higher AHEI-2010 score and lower proportion of ultra-processed foods). In this “no change” cluster, participants were more likely to have kept working outside home during the lockdown period or not to have a professional activity before the lockdown (i.e., unemployed, housemaker, retired). They were also less likely to live in the Paris Region or the East, i.e., regional areas where the epidemic was the most active (Table 2). Cluster 2 (37.4%) corresponded to participants who reported unfavourable nutritional changes or behaviours during the lockdown period: increased weight, decreased physical activity and increased sedentary time, trouble keeping a regular mealtime schedule, buying less fresh products, snacking more than once a day, eating more out of boredom and anxiety, increase in consumption of biscuits/cakes, sweets/chocolate, sandwiches/pizza/savoury pies, potatoes, cheese, and alcoholic drinks, and decrease for fresh products (fruit, vegetables, meat), difficulties to shop in usual stores, or for usual food products (including organic food supplies), stress due to lacking some foods and storage of food products. In this cluster, participants also reported spending more time cooking and that their diet quality did not change. This cluster was composed of participants more likely to be younger, women, non-smokers, with a higher level of education, lower incomes, with children aged under 18 y at home, with higher GAD-7 score for anxiety and PHQ-9 score for depressive symptoms and with a higher proportion of ultra-processed foods in their usual diet. In this “unfavourable nutritional changes” cluster, participants were more likely to work from home during the lockdown period (Table 2). Cluster 3 (19.8%) corresponded to participants who reported favourable nutritional changes or behaviours during the lockdown period: qualitative decrease in consumption reported for biscuits/cakes, sweets/chocolate, sandwiches/pizza/savoury pies and alcoholic drinks, and increase for fresh fruit and vegetables and fish, a diet quality judged better, avoiding some foods or drinks for weight management, willingness to balance the diet, spending more time cooking. This cluster was composed of participants more likely to be younger, overweight/obese, smokers, with a higher level of education and higher incomes, no children aged under 18 y at home, with a higher GAD-7 score for anxiety and a lower PHQ-9 score for depressive symptoms, and with a lower usual diet quality (lower AHEI-2010 score). In this “favourable nutritional changes” cluster, participants were more likely to be partially unemployed or students but also to telework from home during the lockdown period (Table 2).

**Table 2.**
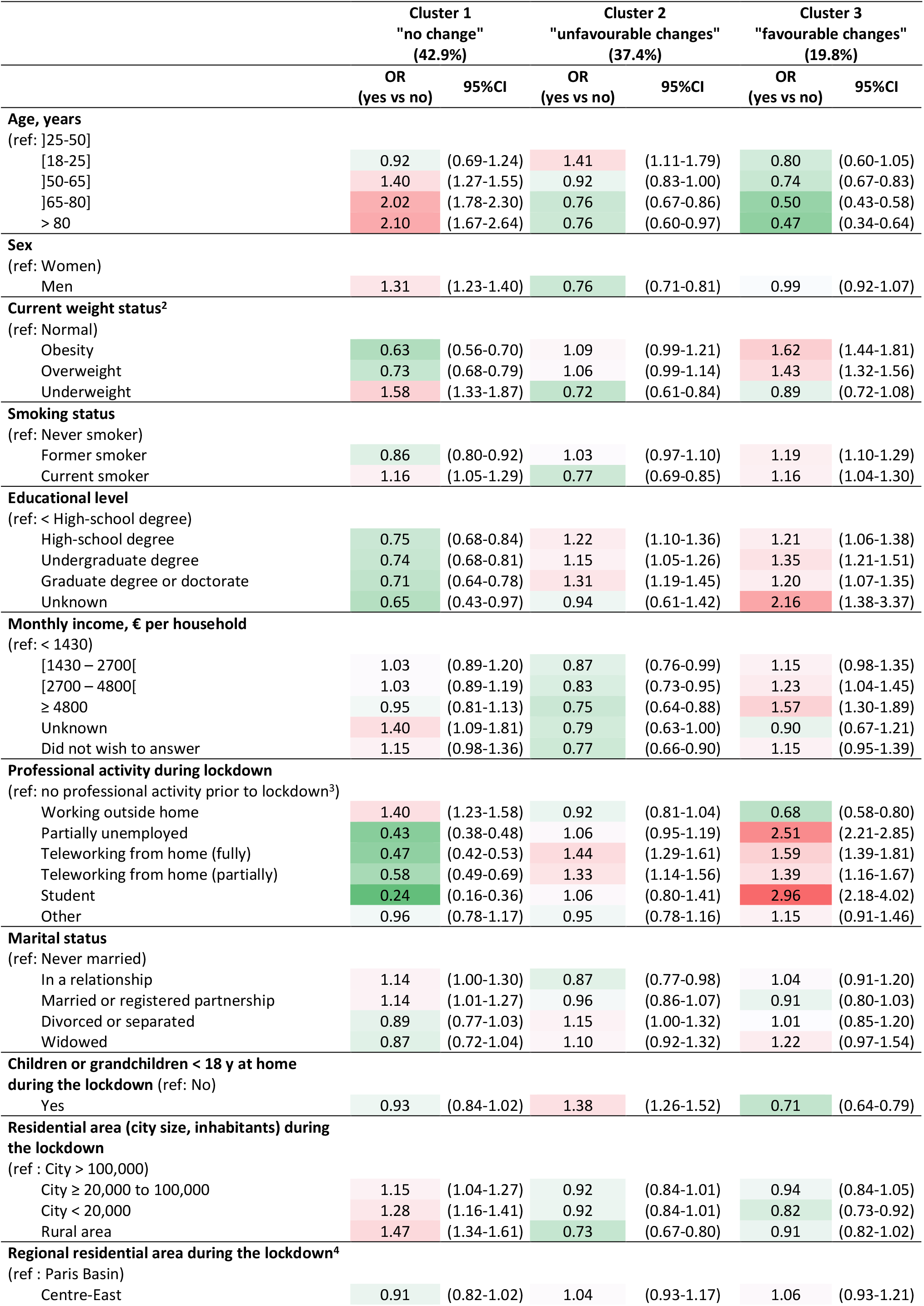
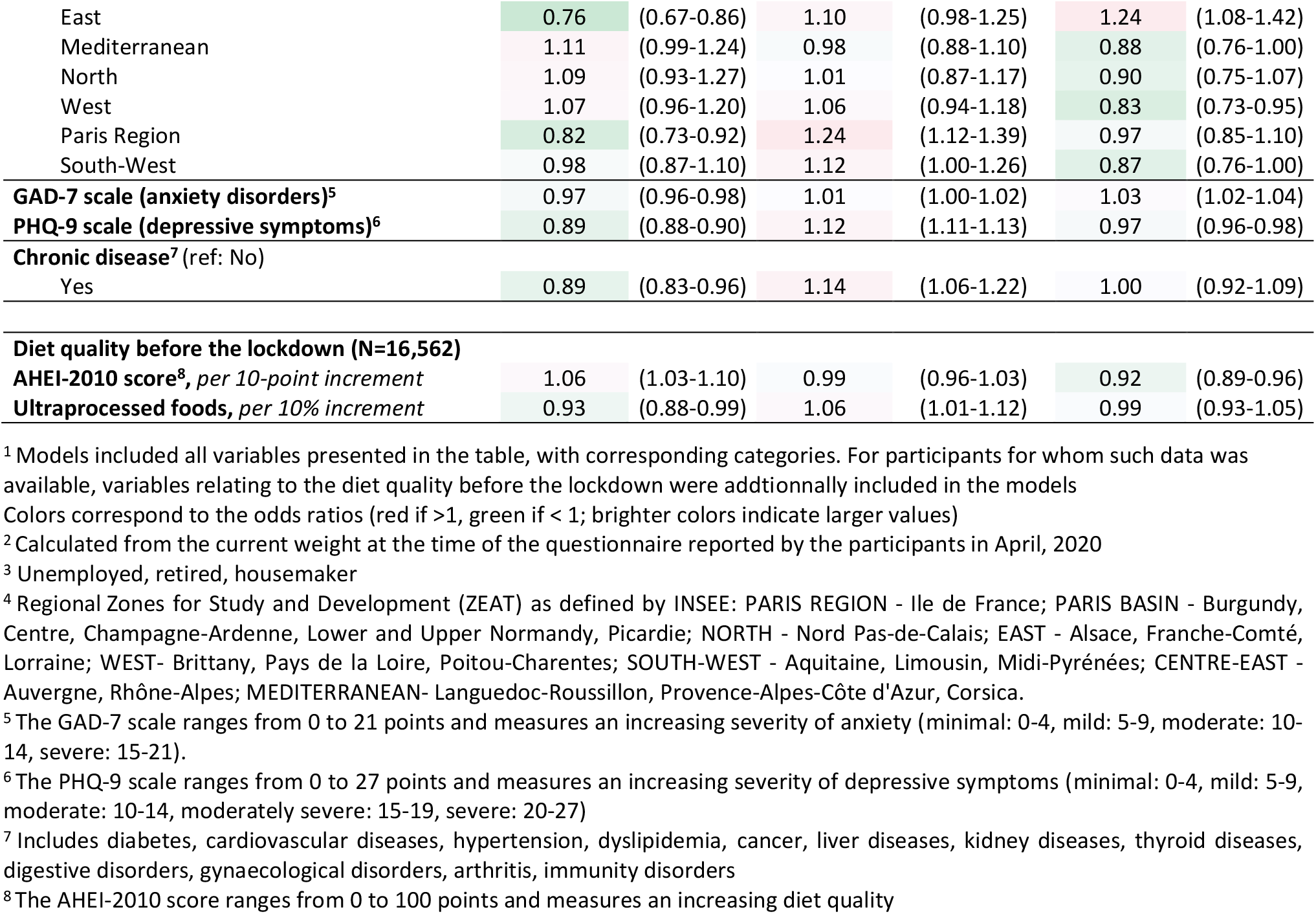
Individual characteristics of participants belonging to each cluster of nutritional behaviours during the lockdown period, multivariable logistic regression models^1^ (N = 22,042)

|  | Cluster 1<br>"no change"<br>(42.9%) |  | Cluster 2<br>"unfavourable changes"<br>(37.4%) |  | Cluster 3<br>"favourable changes"<br>(19.8%) |  |
| --- | --- | --- | --- | --- | --- | --- |
|  | OR<br>(yes vs no) | 95%CI | OR<br>(yes vs no) | 95%CI | OR<br>(yes vs no) | 95%CI |
| **Age, years** |  |  |  |  |  |  |
| (ref: ]25-50] |  |  |  |  |  |  |
| [18-25] | 0.92 | (0.69-1.24) | 1.41 | (1.11-1.79) | 0.80 | (0.60-1.05) |
| ]50-65] | 1.40 | (1.27-1.55) | 0.92 | (0.83-1.00) | 0.74 | (0.67-0.83) |
| ]65-80] | 2.02 | (1.78-2.30) | 0.76 | (0.67-0.86) | 0.50 | (0.43-0.58) |
| > 80 | 2.10 | (1.67-2.64) | 0.76 | (0.60-0.97) | 0.47 | (0.34-0.64) |
| **Sex** |  |  |  |  |  |  |
| (ref: Women) |  |  |  |  |  |  |
| Men | 1.31 | (1.23-1.40) | 0.76 | (0.71-0.81) | 0.99 | (0.92-1.07) |
| **Current weight status<sup>2</sup>** |  |  |  |  |  |  |
| (ref: Normal) |  |  |  |  |  |  |
| Obesity | 0.63 | (0.56-0.70) | 1.09 | (0.99-1.21) | 1.62 | (1.44-1.81) |
| Overweight | 0.73 | (0.68-0.79) | 1.06 | (0.99-1.14) | 1.43 | (1.32-1.56) |
| Underweight | 1.58 | (1.33-1.87) | 0.72 | (0.61-0.84) | 0.89 | (0.72-1.08) |
| **Smoking status** |  |  |  |  |  |  |
| (ref: Never smoker) |  |  |  |  |  |  |
| Former smoker | 0.86 | (0.80-0.92) | 1.03 | (0.97-1.10) | 1.19 | (1.10-1.29) |
| Current smoker | 1.16 | (1.05-1.29) | 0.77 | (0.69-0.85) | 1.16 | (1.04-1.30) |
| **Educational level** |  |  |  |  |  |  |
| (ref: < High-school degree) |  |  |  |  |  |  |
| High-school degree | 0.75 | (0.68-0.84) | 1.22 | (1.10-1.36) | 1.21 | (1.06-1.38) |
| Undergraduate degree | 0.74 | (0.68-0.81) | 1.15 | (1.05-1.26) | 1.35 | (1.21-1.51) |
| Graduate degree or doctorate | 0.71 | (0.64-0.78) | 1.31 | (1.19-1.45) | 1.20 | (1.07-1.35) |
| Unknown | 0.65 | (0.43-0.97) | 0.94 | (0.61-1.42) | 2.16 | (1.38-3.37) |
| **Monthly income, € per household** |  |  |  |  |  |  |
| (ref: < 1430) |  |  |  |  |  |  |
| ]1430 – 2700[ | 1.03 | (0.89-1.20) | 0.87 | (0.76-0.99) | 1.15 | (0.98-1.35) |
| ]2700 – 4800[ | 1.03 | (0.89-1.19) | 0.83 | (0.73-0.95) | 1.23 | (1.04-1.45) |
| ≥ 4800 | 0.95 | (0.81-1.13) | 0.75 | (0.64-0.88) | 1.57 | (1.30-1.89) |
| Unknown | 1.40 | (1.09-1.81) | 0.79 | (0.63-1.00) | 0.90 | (0.67-1.21) |
| Did not wish to answer | 1.15 | (0.98-1.36) | 0.77 | (0.66-0.90) | 1.15 | (0.95-1.39) |
| **Professional activity during lockdown** |  |  |  |  |  |  |
| (ref: no professional activity prior to lockdown<sup>3</sup>) |  |  |  |  |  |  |
| Working outside home | 1.40 | (1.23-1.58) | 0.92 | (0.81-1.04) | 0.68 | (0.58-0.80) |
| Partially unemployed | 0.43 | (0.38-0.48) | 1.06 | (0.95-1.19) | 2.51 | (2.21-2.85) |
| Teleworking from home (fully) | 0.47 | (0.42-0.53) | 1.44 | (1.29-1.61) | 1.59 | (1.39-1.81) |
| Teleworking from home (partially) | 0.58 | (0.49-0.69) | 1.33 | (1.14-1.56) | 1.39 | (1.16-1.67) |
| Student | 0.24 | (0.16-0.36) | 1.06 | (0.80-1.41) | 2.96 | (2.18-4.02) |
| Other | 0.96 | (0.78-1.17) | 0.95 | (0.78-1.16) | 1.15 | (0.91-1.46) |
| **Marital status** |  |  |  |  |  |  |
| (ref: Never married) |  |  |  |  |  |  |
| In a relationship | 1.14 | (1.00-1.30) | 0.87 | (0.77-0.98) | 1.04 | (0.91-1.20) |
| Married or registered partnership | 1.14 | (1.01-1.27) | 0.96 | (0.86-1.07) | 0.91 | (0.80-1.03) |
| Divorced or separated | 0.89 | (0.77-1.03) | 1.15 | (1.00-1.32) | 1.01 | (0.85-1.20) |
| Widowed | 0.87 | (0.72-1.04) | 1.10 | (0.92-1.32) | 1.22 | (0.97-1.54) |
| **Children or grandchildren < 18 y at home during the lockdown (ref: No)** |  |  |  |  |  |  |
| Yes | 0.93 | (0.84-1.02) | 1.38 | (1.26-1.52) | 0.71 | (0.64-0.79) |
| **Residential area (city size, inhabitants) during the lockdown** |  |  |  |  |  |  |
| (ref : City > 100,000) |  |  |  |  |  |  |
| City ≥ 20,000 to 100,000 | 1.15 | (1.04-1.27) | 0.92 | (0.84-1.01) | 0.94 | (0.84-1.05) |
| City < 20,000 | 1.28 | (1.16-1.41) | 0.92 | (0.84-1.01) | 0.82 | (0.73-0.92) |
| Rural area | 1.47 | (1.34-1.61) | 0.73 | (0.67-0.80) | 0.91 | (0.82-1.02) |
| **Regional residential area during the lockdown<sup>4</sup>** |  |  |  |  |  |  |
| (ref : Paris Basin) |  |  |  |  |  |  |
| Centre-East | 0.91 | (0.82-1.02) | 1.04 | (0.93-1.17) | 1.06 | (0.93-1.21) |
| East | 0.76 | (0.67-0.86) | 1.10 | (0.98-1.25) | 1.24 | (1.08-1.42) |
| Mediterranean | 1.11 | (0.99-1.24) | 0.98 | (0.88-1.10) | 0.88 | (0.76-1.00) |
| North | 1.09 | (0.93-1.27) | 1.01 | (0.87-1.17) | 0.90 | (0.75-1.07) |
| West | 1.07 | (0.96-1.20) | 1.06 | (0.94-1.18) | 0.83 | (0.73-0.95) |
| Paris Region | 0.82 | (0.73-0.92) | 1.24 | (1.12-1.39) | 0.97 | (0.85-1.10) |
| South-West | 0.98 | (0.87-1.10) | 1.12 | (1.00-1.26) | 0.87 | (0.76-1.00) |
| **GAD-7 scale (anxiety disorders)<sup>5</sup>** | 0.97 | (0.96-0.98) | 1.01 | (1.00-1.02) | 1.03 | (1.02-1.04) |
| **PHQ-9 scale (depressive symptoms)<sup>6</sup>** | 0.89 | (0.88-0.90) | 1.12 | (1.11-1.13) | 0.97 | (0.96-0.98) |
| **Chronic disease<sup>7</sup> (ref: No)** |  |  |  |  |  |  |
| Yes | 0.89 | (0.83-0.96) | 1.14 | (1.06-1.22) | 1.00 | (0.92-1.09) |
| **Diet quality before the lockdown (N=16,562)** |  |  |  |  |  |  |
| **AHEI-2010 score<sup>8</sup>, per 10-point increment** | 1.06 | (1.03-1.10) | 0.99 | (0.96-1.03) | 0.92 | (0.89-0.96) |
| **Ultraprocessed foods, per 10% increment** | 0.93 | (0.88-0.99) | 1.06 | (1.01-1.12) | 0.99 | (0.93-1.05) |

<sup>1</sup> Models included all variables presented in the table, with corresponding categories. For participants for whom such data was available, variables relating to the diet quality before the lockdown were additionally included in the models
Colors correspond to the odds ratios (red if >1, green if < 1; brighter colors indicate larger values)
<sup>2</sup> Calculated from the current weight at the time of the questionnaire reported by the participants in April, 2020
<sup>3</sup> Unemployed, retired, housemaker
<sup>4</sup> Regional Zones for Study and Development (ZEAT) as defined by INSEE: PARIS REGION - Ile de France; PARIS BASIN - Burgundy, Centre, Champagne-Ardenne, Lower and Upper Normandy, Picardie; NORTH - Nord Pas-de-Calais; EAST - Alsace, Franche-Comté, Lorraine; WEST- Brittany, Pays de la Loire, Poitou-Charentes; SOUTH-WEST - Aquitaine, Limousin, Midi-Pyrénées; CENTRE-EAST - Auvergne, Rhône-Alpes; MEDITERRANEAN- Languedoc-Roussillon, Provence-Alpes-Côte d'Azur, Corsica.
<sup>5</sup> The GAD-7 scale ranges from 0 to 21 points and measures an increasing severity of anxiety (minimal: 0-4, mild: 5-9, moderate: 10-14, severe: 15-21).
<sup>6</sup> The PHQ-9 scale ranges from 0 to 27 points and measures an increasing severity of depressive symptoms (minimal: 0-4, mild: 5-9, moderate: 10-14, moderately severe: 15-19, severe: 20-27)
<sup>7</sup> Includes diabetes, cardiovascular diseases, hypertension, dyslipidemia, cancer, liver diseases, kidney diseases, thyroid diseases, digestive disorders, gynaecological disorders, arthritis, immunity disorders
<sup>8</sup> The AHEI-2010 score ranges from 0 to 100 points and measures an increasing diet quality

## DISCUSSION

This study, conducted on > 37,000 French adults provide an overview of the nutritional behaviours observed during the lockdown set in the midst of the COVID-19 pandemic to control the spread of the disease.

Our results highlight several trends in nutritional behaviours during the lockdown.

On one hand, there was a trend towards rather unfavourable nutritional behaviours, with increased energy intakes, decreased physical activity levels and increased sedentary time, and eventually weight gain (+1.8kg in two months). The weight gained in this short period of time may become permanent in some individuals and further lead to more weight gain in the future, all the more if the unfavourable nutritional behaviours observed during the lockdown are not reversed [29]. This is of concern, especially since individuals more likely to have gained weight were already overweight or obese at the beginning of the lockdown. Beyond energy balance, we also observed trends towards increasing snacking behaviours (several times a day, every day, when the public health recommendation is to avoid snacking between main meals [28]) and a consistent increase reported for the consumption of sweets, chocolate, biscuits or cakes. Individuals also reported buying fewer fresh products, notably due to less frequent shopping or difficulties in accessing their usual food stores or finding their usual food products (especially for older individuals). Decreasing the consumption of fresh vegetables likely led to increases in frozen or canned vegetables intake, yet a similar compensation did not appear for fruit or for fish, raising the question of substitution between these different types of products and their respective supply. In particular, the lower intake of fish may be of particular concern, since it consistently translated into a reduced intake of related nutrients such as long chain n-3 polyunsaturated fatty acids or iodine. In line, lower intakes of vitamin D were also observed, in a context where vitamin D status is likely already strongly impacted by the lockdown, the majority of vitamin D being produced following sun exposure (which was often not compatible with the requirement to stay home). The lockdown also resulted in reported nutritional behaviours linked to the disruption of habits and notably the loss of activity (work or leisure-related), such as eating to compensate for boredom or anxiety or having trouble keeping regular mealtime schedules. For some individuals, an increase in alcohol intake was also reported. This overall unhealthy pattern of nutritional behaviours may have resulted from the deprivation of opportunities due to the lockdown: loss of options for exercise, food shopping or meals due to the closure of sport facilities, restaurants and workplaces, the restricted access to food supplies and the instructions for adults and children to stay at home. Along with this pattern of unhealthy behaviours, participants also declared that they spent more time cooking, a rather favourable behaviour, but which may have contributed here to the increase consumption of biscuits or cakes. This overall pattern particularly clustered participants (cluster 2) who were women, with a higher level of education but lower income, teleworking from home (which was generalized as a result of the lockdown measures), with children aged under 18 y at home, higher scores for depressive symptoms, and already a higher proportion of ultra-processed foods in their diets before the lockdown. Less healthy nutritional behaviours have also been underlined in studies involving children and students in Italy [30,31] where the lockdown had a negative impact on physical activity but also on the consumption of foods, which is in line with this first trend of behaviours.

On the contrary, we also highlighted a second trend of more favourable nutritional behaviours, where individuals decreased their energy intake, increased or maintained their physical activity and eventually lost weight. Such modifications being made knowingly to control weight or balance the ratio of energy intake/expenditure. Indeed, some individuals also considered this lockdown period as an opportunity to spend more time cooking and to balance their overall diet. Improvements in the nutritional quality of diets were observed: increased AHEI-2010 score for diet quality, decreased proportion of ultra-processed foods, increased intakes of fruit and vegetables, legumes or nuts, decreased intakes of red meat or alcohol. These favourable behaviours mostly clustered participants (cluster 3) who were overweight/obese, smokers, with a higher level of education and income, who were short-time working, students (in this case, more likely students who returned to their parents’ during the lockdown) or teleworking from home, with no children under 18 y at home, and exhibiting more anxiety but less depressive symptoms. Looking at their usual diet before the lockdown, these individuals were those with a lower diet quality. Overall, individuals adopting healthier nutritional behaviours during the lockdown were probably those already more likely to do so or with more potential for improvement. For these individuals, the lockdown, by disrupting habits, might have led to a favourable adaptation, considering and adopting newer options beyond established habits of what they were used to do or buy, or where they usually shopped. It could also be that these individuals were more concerned about their health, notably as regards their risks for COVID-19 infection or prognosis (e.g., overweight or obese individuals).

Finally, a third group reported stable nutritional behaviours during the lockdown period. These clustered participants (cluster 1) who were older, living in rural areas, with no professional activity before the lockdown period (i.e., unemployed, housemaker, retired) or those who kept working outside home. Hence, corresponding to individuals with probably less disruption in their lifestyle or environment or with more settled habits.

The experience of lockdown may have very different forms and related consequences when considering socioeconomic inequalities. Our results suggested that higher incomes were associated with more favourable modifications of nutritional behaviours (individually and combined as cluster 3): spending more time cooking, intentional reduction of food intake to avoid weight gain or compensate for lower physical activity, healthy changes in food consumption. These individuals were most likely those with more means and options to undertake modifications. On the other hand, lower incomes associated with less favourable modifications of nutritional behaviours (individually and combined as cluster 2): eating in response to boredom or anxiety, trouble keeping regular mealtime schedules, stress of lacking some food during lockdown, buying fewer fresh products, unhealthy changes in food consumption. Individuals with a higher level of education presented favourable nutritional behaviour changes as reflected in cluster 3 (especially spending more time cooking), but also less favourable changes reflected in cluster 2 (especially snacking). On the other hand, individuals with a lower educational level exhibited a mixed profile: more likely to belong to the “no change” cluster 1 but with trouble keeping regular mealtime schedule, buying fewer fresh products but also adjusting dietary intakes to their level of exercise. Hence, the level of education and income did not necessarily correspond to nutritional behaviours and changes in the same way. This is likely related to the professional activity during lockdown, with most individuals forced to stay home (which induced lifestyle changes). Individuals who kept working at home (teleworking, most often corresponding to more qualified jobs) were present in clusters 2 and 3, suggesting opposite directions for nutritional behaviours, which may be related to their experience during the lockdown (e.g., time spent working, living with others, presence of children at home). In particular, the presence of children at home which associated with cluster 2 may be linked to more snacking and to the subsequent consumption of sweets, biscuits and cakes. Other individuals became partially unemployed, and were more likely to adopt favourable nutritional behaviours (cluster 3). Finally, those who kept working outside of their home (essential activities; often less qualified jobs, except for those working in medical care) or those with no professional activity before the lockdown period (unemployed, retired, housewife or husband) generally exhibited few modifications in nutritional behaviours compared to others (cluster 1).

Strengths of our study pertained to its large sample size and the in-depth characterisation of participants, including nutritional behaviours throughout follow-up and more specifically during the lockdown period, with qualitative and also quantitative assessments. The flexibility of the web platform of the NutriNet-Santé study allowed to quickly implement lockdown-related studies (development, assessment and treatment/analysis) to provide insights into multiple nutritional aspects of this singular situation at the large-scale level. However, some limitations should be acknowledged. Indeed, the NutriNet-Santé study is a long-term prospective study focusing on nutrition and health and based on continued voluntary recruitment since 2009. This implies that the NutriNet-Santé population is composed of more women and individuals with overall higher socioeconomic position compared to the general French population [32,33]. All the analyses were weighted to improve representativeness to overcome this bias. Still, our study likely does not accurately capture the experience of more disadvantaged or older populations. Besides, NutriNet-Santé participants show an interest in nutrition and health and may therefore adopt healthier nutritional behaviours than the overall French population and to consciously rethink their nutritional behaviours to adapt to the new situation and make the most out of it. Still, trends of unfavourable nutritional behaviours during lockdown were observed in this more “health-conscious” population, which raises even more concerns for the general population.

To conclude, our results suggested that the lockdown led, in a substantial part of the population, to unhealthy nutritional behaviours that, if maintained in the long term, may increase the nutrition-related burden of disease [3]. A particular vigilance will therefore be needed to assess whether individuals return to healthier nutritional habits at the end of the COVID-19 pandemic, especially considering that unfavourable nutritional behaviours were more likely to be observed in individuals who worked from home during the period of strict lockdown (and are still encouraged to keep doing so afterwards). Nonetheless, our study also revealed that the lockdown situation also created, at least in some (non-negligible) segments of the population, an opportunity to improve nutritional behaviours, such as cooking homemade meals, increasing consumption of fresh products and buying food products from local shop and/or farmers. Understanding the leverages to put these healthier nutritional behaviours on a long-term footing (e.g., time, tools, food supply at work, local food supply) post-lockdown, and try to make the most out of this momentum, will be key to improving nutritional behaviours at a broader scale in the future. This first descriptive study gave us many clues of how the COVID-19 crisis has affected nutritional behaviours. Further studies are required to better understand the mechanisms behind these changes. Beyond this report, the data collected in the framework of the SAPRIS project, combined with the extensive nutritional characterization of participants in the NutriNet-Santé study, will also allow to study how nutrition associates with the risk of developing COVID-19 (symptoms and serology that will be soon available for tens of thousands of participants).

## Data Availability

Data described in the manuscript (limited to summary data per participant consent), code book, and analytic code will be made available upon reasonable request pending application and approval.

## ACKNOWLEDGEMENTS

The authors warmly thank all the volunteers of the NutriNet-Santé cohort for their continuous participation in the study and for participating in this COVID-19-specific project. The author thank the SAPRIS project working group (management board: Dr Nathalie Bajos co-PI, Dr Fabrice Carrat, co-PI, Dr Marie Zins, Dr Gianluca Severi, Dr Marie-Aline Charles, Dr Pierre-Yves Ancel, Dr Mathilde Touvier). We also thank Thi Hong Van Duong, Régis Gatibelza, Jagatjit Mohinder, and Aladi Timera (computer scientists); Nathalie Arnault, Julien Allegre, and Laurent Bourhis (datamanager/statisticians); Cédric Agaesse (dietitian); Fatoumata Diallo, Roland Andrianasolo, and Sandrine Kamdem (physicians) for their technical contribution to the NutriNet-Santé study.

